# Cross-Device Adaptation of Mirai for Mammography-Based Breast Cancer Risk Prediction

**DOI:** 10.64898/2026.06.15.26355696

**Authors:** Adriana Sistig, Joseph H. Rothstein, Tejomay Gadgil, Ninah Achacoso, Stacey E. Alexeeff, Lawrence D. Gerstley, Robert J. Klein, Laurie R. Margolies, Albert Pu, Cara L. Smith Gueye, Marvella Villaseñor, Mark Westley, Laurel A. Habel, Vignesh A. Arasu, Weiva Sieh, Li Shen

## Abstract

Fine-tuning can adapt pretrained medical imaging models to new clinical datasets, but device-specific domain shifts may limit generalizability. We evaluated Mirai, a mammography-based deep learning model for breast cancer risk prediction, in a large screening cohort containing Hologic and General Electric (GE) full-field digital mammography systems, including GE Premium View (GE PV) and Tissue Equalization (GE TE) post-processing software. Native Mirai showed lower performance on TE images than on Hologic or PV images. Fine-tuning on TE images improved TE performance, particularly for short-term risk prediction, but substantially reduced performance on Hologic images, consistent with catastrophic forgetting. To mitigate this effect, we developed a device-invariant model using interleaved multi-device sampling and conditional adversarial training. This approach largely restored Hologic performance while maintaining improved TE performance, providing better robustness across heterogeneous imaging platforms. Comparison of cumulative and annual risk AUCs over a five-year time horizon further showed that performance gains were driven mainly by short- and intermediate-term predictions. These findings highlight both the value and dangers of device-specific fine-tuning and support balanced domain-adaptation strategies for deploying mammography-based risk models across diverse clinical imaging environments.

## Introduction

Mammography-based deep learning models have shown promise for breast cancer (BC) risk prediction, with Mirai, developed by Yala et al. (2021), demonstrating stronger performance than traditional clinical risk models such as Gail and Tyrer–Cuzick (1-3). However, Mirai was developed using full-field digital mammography images acquired on Hologic systems, and its performance may be affected when applied to images from other manufacturers or post-processing pipelines (1, 4, 5).

This limitation is clinically important because real-world screening programs often use heterogeneous imaging platforms (1, 6, 7). Differences in manufacturer, detector characteristics, and post-processing software can shift image appearance and degrade model generalizability (8–12). Although fine-tuning is a practical strategy for adapting pretrained models to new imaging domains, device-specific fine-tuning may also introduce bias toward the target device and reduce performance on the original domain, a phenomenon referred to as catastrophic forgetting (13).

In this study, we evaluated Mirai in a large Kaiser Permanente Northern California (KPNC) screening mammography cohort that included Hologic and General Electric (GE) systems, including two GE post-processing software types: Premium View (GE PV) and Tissue Equalization (GE TE). We then fine-tuned Mirai on the previously unseen GE domain and evaluated whether interleaved multi-device sampling and conditional adversarial training (CAT) could improve cross-device robustness while preserving performance gains on both Hologic and GE images.

## Methods

### Study Cohort Description

The study population comprised 178,000 women with screening full-field digital mammography (FFDM) examinations at KPNC between 2003 and 2020 (1). Specifically examinations with Breast Imaging Reporting and Data System (BI-RADS) category 1 or 2, or a screening BI-RADS 0 and diagnostic BI-RADS 1 or 2 in 90 days or less, or a screening BI-RADS 0 and diagnostic BI-RADS 4 or 5 and radiologic-pathologic concordant benign biopsy in 90 days or less (1). FFDM images were acquired on Hologic (2004–2020) or GE machines. GE images were processed with either Premium View (2006–2020) or Tissue Equalization (2003–2016) post-processing software. PV and TE were determined using the Acquisition Device Processing Code DICOM tag, “PREMIUM_VIEW” or “Proc_1” respectively. The PV device was developed by GE Medical Systems to improve diagnostic confidence by enhancing contrast to increase the visibility of malignancies and introduced into KPNC in 2006 (14, 15). The TE device is an older post-processing software that does not provide local contrast enhancement, and instead applies tissue equalization to balance the contrast and brightness of an image to ensure that adipose and fibroglandular areas are both clearly visible (7, 15). Given the visual difference between PV and TE, they are treated as two separate devices in this study. Participants were excluded if they had: a prior history of BC or breast implants. Additionally, participants were excluded if any of the four standard screening mammography views, left and right craniocaudal (CC) and mediolateral oblique (MLO), were missing. After these exclusions, a total of 152,165 unique patient exams comprising 115,774 Hologic, 11,866 GE PV, and 24,525 GE TE remained for analysis (Table 1).

**Table 1.** Distribution of time to diagnosis of breast cancer across Hologic, GE Premium View, and GE Tissue Equalization devices.

| Time to Breast Cancer<br>(Years) | Hologic | General Electric |  | Total |
| --- | --- | --- | --- | --- |
|  |  | Premium View | Tissue<br>Equalization |  |
| 0-1 | 1037 | 103 | 183 | 1323 |
| 1-2 | 457 | 54 | 121 | 632 |
| 2-3 | 494 | 55 | 86 | 635 |
| 3-4 | 434 | 45 | 102 | 581 |
| 4-5 | 459 | 52 | 79 | 590 |
| No cancer diagnosed<br>within 5 years | 112893 | 11557 | 23954 | 148404 |
| **Total** | **115774** | **11866** | **24525** | **152165** |

### Methodology Overview

We evaluated performance of the original (native) Mirai software across the full cohort and separately by device domain. We then fine-tuned Mirai using GE TE examinations to adapt the model to this previously unseen domain. Model development and evaluation used 5-fold cross-validation (CV), with training, development, and test partitions stratified by time to cancer and device type. Performance was assessed using Mirai’s time-dependent inverse-probability-of-censoring-weighted (IPCW) concordance index (C-index) as the overall 5-year discrimination metric, and area under the receiver operating characteristic curve (AUC) for specific time intervals.

Three model configurations were compared: 0) native Mirai (“Native”), 1) TE-specific fine-tuned Mirai (“Finetuned”), and 2) a device-invariant model (“Device Invariant”). Evaluation of Mirai across TE, PV, and Hologic machines revealed a statistically significant difference between TE and Hologic, but not between PV vs. Hologic. Therefore a fine-tuned model was designed to improve performance specifically on GE TE images. The device-invariant model was initialized from the TE fine-tuned model and further trained using interleaved multi-device sampling to balance the representation of Hologic, GE PV, and GE TE images. Conditional adversarial training (CAT) was also used to encourage device-agnostic image representations while preserving cancer risk prediction performance.

Confidence intervals were estimated using 5,000 bootstrap samples. To compare models, pairwise differences in C-index were evaluated using paired bootstrap resampling, with statistical significance inferred when the 95% confidence interval of the difference excluded zero. Statistical significance of pairwise differences in model AUCs for specific time intervals was assessed using DeLong’s nonparametric test for correlated ROC curves, with differences considered statistically significant when the *P*-value was < 0.0056, applying a Bonferroni correction for the three models and three device types evaluated (16, 17). Additional implementation details, including batch normalization momentum adjustment, interleaved sampling, CAT, and cumulative versus annual risk AUC definitions, are provided in the Supplementary Methods.

## Results

Evaluation on the full cohort (Figure 1) showed that the native Mirai had a lower overall C-index for TE (0.71; 95% CI: 0.69–0.73) than for Hologic (0.74; 0.73–0.75) or PV (0.74; 0.71–0.77). Further analysis of annual risk AUC scores highlighted Mirai’s stronger performance in predicting short-term risk compared with long-term risk (Supplementary Figure 1). Additionally, the reduced performance on TE devices was mainly driven by short-term (0-2 years) risk prediction. The annual risk AUC for Hologic vs. TE was 0.87 (0.85-0.88) vs. 0.81 (0.78–0.84) in the first year, and 0.71 (0.69–0.73) vs. 0.64 (0.59–0.69) in the second year.

**Figure 1.**
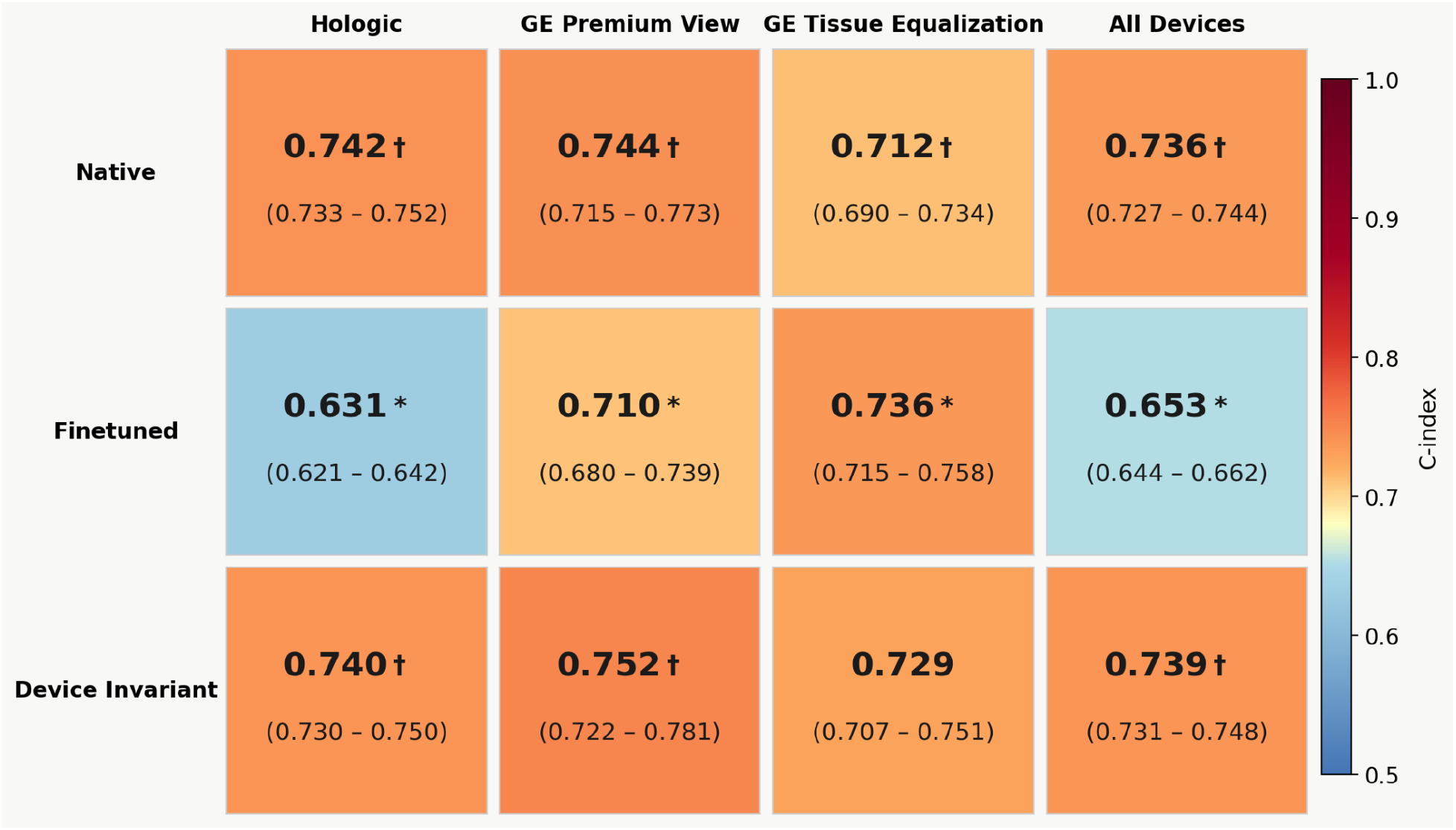
Overall 5-year discrimination assessed using Mirai’s time-dependent inverse-probability-of-censoring-weighted (IPCW) concordance index (C-index) in a cohort of 152,165 women, by model and imaging device. 95% confidence intervals were estimated using 5,000 bootstrap resamples. Pairwise model comparisons were performed using paired bootstrap testing with 5,000 resamples. Statistical significance was defined as a 95% bootstrap confidence interval for the paired difference that excluded 0. * indicates a statistically significant difference from the Native model for the corresponding device † indicates a statistically significant difference from the Finetuned model for the corresponding device.

Fine-tuning the native Mirai model using only GE TE devices, improved risk prediction on GE TE over the 1–5-year horizon, with statistically significant gains in both the C-index and per-year AUCs compared with Native (Figure 1 and Supplementary Figure 1). However, the Fintuned model performance on Hologic devices declined significantly, with the C-index decreasing from 0.74 (0.73–0.75) to 0.64 (0.63–0.65) reflecting catastrophic forgetting (13). The fine-tuned model performance on PV also decreased although the difference was not statistically significant. These findings suggest that fine-tuning on TE only led to device-specific performance gain at the expense of generalizability to other devices, motivating the development of a device-invariant model.

To mitigate the risk of catastrophic forgetting, a device-invariant was developed from the fine-tuned model by reintroducing the Hologic and PV devices via continued learning using an interleaved data sampler to balance all the devices during training. Furthermore, CAT was used to force the model to generate device-invariant features. The device-invariant model’s C-indices on Hologic and PV recovered to a level comparable to native Mirai, while maintaining similar performance to the TE fine-tuned model (Figure 1 and Supplementary Figure 1). These results show that although the device-specific fine-tuning achieved the highest performance on TE, the device-invariant model demonstrated improved cross-device generalizability and overall best performance among the three models. Additionally, Supplementary Figure 1 shows that most of the model differences were found for short to intermediate risk prediction within the first 3 years.

## Discussion

Using a large heterogeneous screening mammography cohort, we found that fine-tuning Mirai on GE TE images improved performance on this previously unseen device type, particularly for short-term breast cancer risk prediction. However, TE-only fine-tuning substantially reduced performance on Hologic images, consistent with catastrophic forgetting. This finding highlights an important unintended consequence of adapting medical imaging AI models to a new domain: device-specific gains may come at the expense of generalizability.

The device-invariant model using interleaved sampling and CAT largely restored Hologic and PV performance while maintaining improved TE performance. Although the TE-specific model achieved the highest performance on TE images, the device-invariant model provided more robust performance across devices. These results suggest that balanced multi-device training is important when deploying risk models in heterogeneous clinical settings.

Our findings also indicate that image post-processing software, not only manufacturer, influences model behavior. Although both PV and TE images were acquired on GE systems, PV performance more closely resembled Hologic than TE. This may reflect the contrast-enhancing effects of PV processing, which produces images that are visually more similar to those from Hologic systems.

Finally, comparison of cumulative and annual risk AUCs showed that long-term improvements should be interpreted cautiously. Under the cumulative risk definition, early cancer cases are counted as positives when assessing longer time horizons, which can inflate the apparent long-term performance even when there is no improvement in later years. Annual risk prediction performance suggested that benefits of fine-tuning were concentrated mainly in short-to intermediate-term risk prediction.

Overall, this study demonstrates both the value and dangers of fine-tuning mammography-based risk models. Device-specific fine-tuning may be useful for single-platform deployment, but multi-device clinical environments require adaptation strategies that preserve performance across imaging devices.

## Data Availability

Data used in this study were provided by the Kaiser Permanente Research Bank (KPRB) from the KPRB collection, which includes the Kaiser Permanente Research Program on Genes, Environment, and Health (RPGEH). Access to data used in this study may be obtained by application to the KPRB at kp.org/researchbank/researchers.

## Acknowledgements

We thank the participating Kaiser Permanente women, facilities, and medical providers for the data they have provided for this work. This work was funded by the National Cancer Institute (R01CA264987, P01CA281826). This work was supported in part through the Minerva computational and data resources and staff expertise provided by Scientific Computing and Data at the Icahn School of Medicine at Mount Sinai and supported by the Clinical and Translational Science Awards (CTSA) grant UL1TR004419 from the National Center for Advancing Translational Sciences. Research reported in this publication was also supported by the Office of Research Infrastructure of the National Institutes of Health under award number S10OD030463 and S10OD038231. The content is solely the responsibility of the authors and does not necessarily represent the official views of the National Institutes of Health.

