## Supplementary Methods for "Cross-Device Adaptation of Mirai for Mammography-Based Breast Cancer Risk Prediction"

### Study Cohort Collection Procedures

A Current Procedural Terminology examination code of 77057 was used to identify screening FFDM exams. Incident BC, detected either symptomatically or on a subsequent mammogram, was defined as pathology-confirmed invasive carcinoma or ductal carcinoma in situ. Women were followed from their initial index screening FFDM exam date to their BC diagnosis, death, health plan disenrollment (allowing up to a 3-month gap in enrollment) date, or, if none of the previous were noted, the end of the study period (August 31, 2021), whichever occurred first.

### Model Training & Evaluation

Mirai was implemented using the GitHub version accessed September 2023 (Mirai; https://github.com/yala/Mirai). A 5-fold cross validation (CV) strategy was employed for model fine-tuning and evaluation. For each fold, the dataset was partitioned into training, development, and test sets in approximately 60:20:20 proportions, stratified by years to cancer and device type. Each model was trained for 10 epochs with the optimal checkpoint determined by performance (native Mirai C-Index) on the development set, and subsequently evaluated on the held-out test set. The test set predictions across the 5 folds were aggregated for evaluation. We followed the released Mirai evaluation implementation. The C-index was calculated using Mirai's time-dependent IPCW concordance metric based on predicted cumulative risk estimates across the 1–5-year prediction horizon. AUC analyses were performed for both cumulative and annual risk. For annual risk, each cancer case contributed only to the specific follow-up year in which the diagnosis occurred.

The performance of native Mirai was evaluated on the full dataset to establish a baseline. The confidence intervals of C-indices for individual models were estimated using 5,000 bootstrap samples. To compare models, the pairwise differences of the respective C-indices within each bootstrap sample were used to compute the 95% confidence intervals. A 95% confidence interval that excluded 0 was considered a statistically significant difference. Additionally, differences between model AUCs computed for specific time intervals were compared using DeLong's test, implemented using the roc_comparison GitHub repository (https://github.com/yandexdataschool/roc_comparison) developed by Sun & Xu (2014) (17). The DeLong test is a nonparametric method for comparing correlated ROC curves derived from the same study population. A two-sided $P$ value < 0.0056 applying a Bonferroni correction for the three models and three device types compared was considered statistically significant.

### Domain adaptation via Batch Normalization (BN)

BN is widely used to improve the training dynamics of deep neural networks. Mirai uses a residual network as an image encoder, which contains a BN layer for every convolutional layer. However, large shifts in BN running statistics can trigger a cascade of sudden parameter updates, potentially damaging pretrained representations. Supplementary Figure 2 illustrates the differences between Mirai's pretrained statistics and the statistics derived from Hologic and TE in this study. While the Hologic statistics are similar to Mirai's pretrained statistics since Mirai was trained on Hologic images, the TE statistics show significant differences, especially in variances.

To alleviate the impact of different BN statistics between two devices during training, we adjusted the momentum factor for BN (0.1, 0.01, and 0.001) as a hyperparameter and found 0.01 to provide the best performance (19, 20). This is lower than the default value of 0.1, which allows a model to adapt more slowly to the novel distribution thereby improving the fine-tuning results.

### Interleaved sampling for continual learning

Previous work by Chen, et al., has highlighted the challenge of catastrophic forgetting: it occurs when a network is trained sequentially on multiple tasks (e.g., transferring a model trained on Hologic to GE), as the model's parameters are updated to satisfy the objectives of the new task, its performance degrades on the original task (14). To address this issue, we used a continual learning strategy that continues to train a model after it has finished fine-tuning on a new device. It works by applying an interleaved sampler that generates batches of samples each of which comes from a single device in a round-robin fashion, ensuring all devices are equally represented during continual learning. The goal of this method was to create a model that generalizes across multiple devices (18).

**Device-invariant representation learning**

To encourage a model to learn device-agnostic features for cancer risk prediction, conditional adversarial training (CAT) is employed to minimize the image encoder's ability to predict the device type, while at the same time maximize its ability to predict cancer risk. CAT was originally used in Mirai to homogenize the features extracted from two different Hologic device types, Hologic Selenia and Selenia Dimensions. We extended CAT to the three devices in our cohort (Hologic, GE PV, and GE TE) to make the models more generalizable than the models trained with fine-tuning only.

**Cumulative vs. annual risk prediction**

To evaluate Mirai's performance, we included annual risk (1-year intervals) alongside Mirai's cumulative risk approach. Using the cumulative definition, for each time interval $i$, positive cases were women diagnosed with breast cancer within 0 to $i$ years of the index mammogram, and negative controls were women who remained cancer-free with at least $i$ years of follow-up (years to last follow-up $\geq i$ and years to cancer $\geq i$) (Supplementary Table 1). In the annual risk definition, for any given year, a patient is considered positive only if they develop cancer within that specific 1-year window following the index mammogram. Specifically, the negative control criteria were identical; however, positive cases were restricted to women diagnosed with breast cancer exactly in the time period from year $i-1$ to year $i$, and women ultimately diagnosed with cancer whose diagnosis had not yet occurred contributed as negative controls to earlier year windows, ensuring each woman contributed as a positive case to only one 1-year window (Supplementary Table 2). Cumulative and annual AUC's were calculated, for each time interval, using sklearn v1.4.2 (roc_auc_score) function.

We found that the cumulative risk estimation method, while consistent with Mirai's implementation, may yield inflated performance estimates because BC cases are included as positive cases for all subsequent years, effectively assessing whether cancer occurs within all years evaluated. In contrast, the annual risk approach evaluates discrimination within distinct 1-year windows, providing a more temporally precise and conservative assessment of model performance over the 5-year time horizon.

In the evaluation of annual risk prediction, fine-tuning of the TE model resulted in a statistically significant improvement over native Mirai's performance only during the first year (Supplementary Figure 1, Supplementary Table 2). In contrast, the evaluation of cumulative risk prediction demonstrated significant improvement across all 5 years (Supplementary Figure 1 and Supplementary Table 1). These findings suggest that the performance gains observed under the cumulative risk evaluation are largely driven by detection of breast cancers present on the screening mammogram and highlight the TE fine-tuned model's improvement only in short-term risk prediction. Incorporating this knowledge into future analyses of Mirai may provide a more comprehensive understanding of potential performance gains. For this study, including both cumulative and annual risk assessments enabled a more comprehensive and clinically relevant evaluation of predictive performance across a 5-year time horizon.

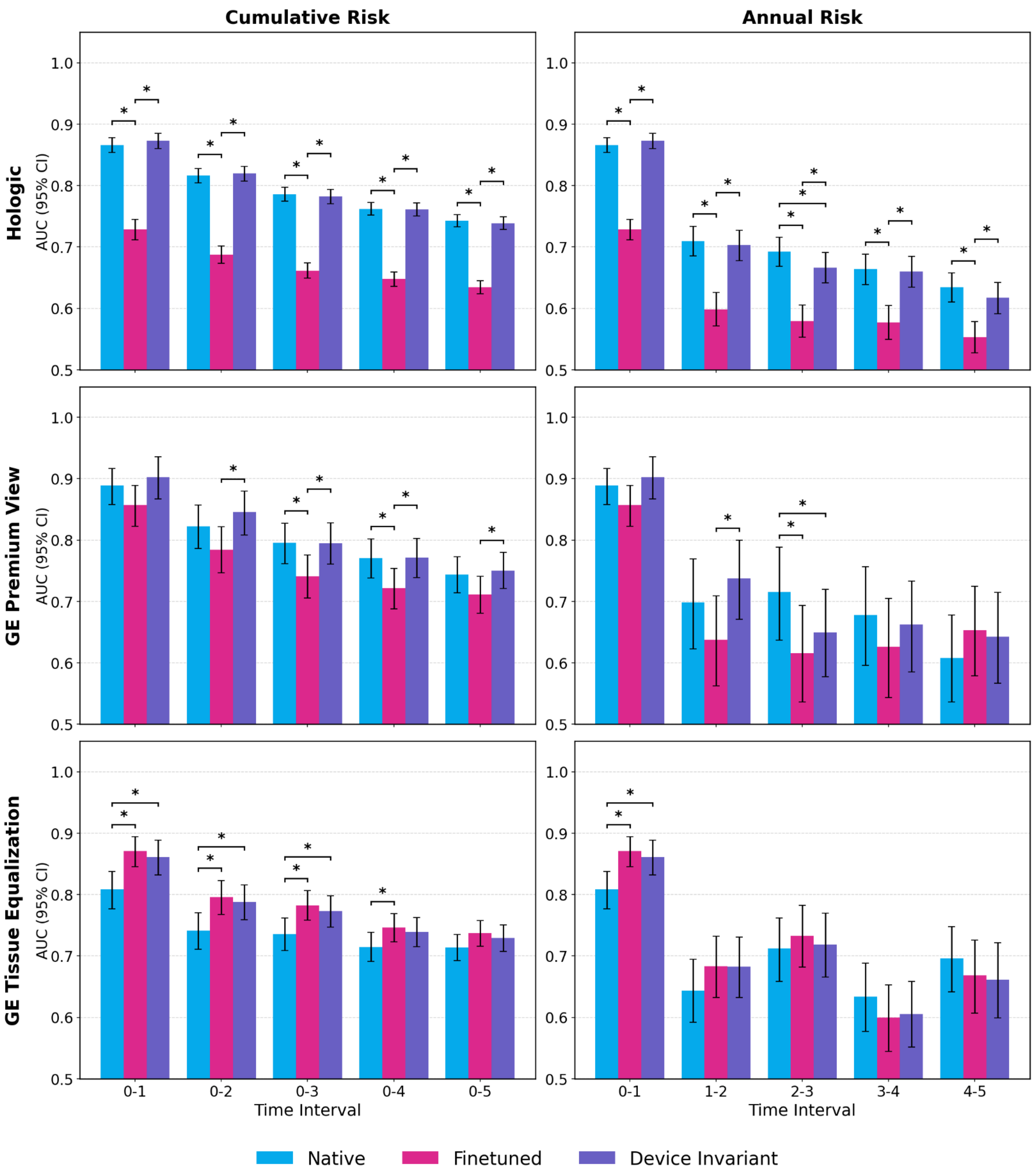


**Supplementary Figure 1.** Discriminatory accuracy (AUC and 95% CI) for cumulative (left) and annual (right) risk predictions of the native Mirai (Native), fine-tuned model for GE Tissue Equalization (Finetuned), and device-invariant model (Device Invariant), by device type. Asterisks (*) denote statistical significance based on Bonferroni-adjusted threshold ($p < 0.0056$) for the three models and three device types compared.

| Cumulative Risk Time Interval | Number of cases/controls | Native Mirai AUC (95% CI) | TE Finetuned AUC (95% CI) | Device Invariant AUC (95% CI) | Finetuned vs. Native (p-value) | Device Invariant vs. Native (p-value) | Device Invariant vs. Finetuned (p-value) |
|---|---|---|---|---|---|---|---|
| **Hologic** | | | | | | | |
| 0-1 | 115774 (1037/114737) | 0.87 (0.85, 0.88) | 0.73 (0.71, 0.74) | 0.87 (0.86, 0.89) | **9.7E-68** | 1.6E-01 | **1.8E-74** |
| 0-2 | 113043 (1494/111549) | 0.82 (0.80, 0.83) | 0.69 (0.67, 0.70) | 0.82 (0.81, 0.83) | **1.7E-78** | 4.1E-01 | **1.5E-85** |
| 0-3 | 110123 (1988/108135) | 0.79 (0.78, 0.80) | 0.66 (0.65, 0.67) | 0.78 (0.77, 0.79) | **6.3E-93** | 3.3E-01 | **8.1E-90** |
| 0-4 | 106394 (2422/103972) | 0.76 (0.75, 0.77) | 0.65 (0.64, 0.66) | 0.76 (0.75, 0.77) | **2.3E-97** | 8.0E-01 | **3.0E-95** |
| 0-5 | 102239 (2881/99358) | 0.74 (0.73, 0.75) | 0.63 (0.62, 0.65) | 0.74 (0.73, 0.75) | **8.2E-101** | 2.5E-01 | **1.5E-90** |
| **GE Premium View** | | | | | | | |
| 0-1 | 11866 (103/11763) | 0.89 (0.86, 0.92) | 0.86 (0.82, 0.89) | 0.90 (0.87, 0.94) | 3.8E-02 | 4.3E-01 | 1.6E-02 |
| 0-2 | 11624 (157/11467) | 0.82 (0.79, 0.86) | 0.78 (0.75, 0.82) | 0.85 (0.81, 0.88) | 8.9E-03 | 8.4E-02 | **1.4E-04** |
| 0-3 | 11393 (212/11181) | 0.80 (0.76, 0.83) | 0.74 (0.70, 0.78) | 0.80 (0.76, 0.83) | **3.1E-05** | 9.7E-01 | **8.9E-05** |
| 0-4 | 11105 (257/10848) | 0.77 (0.74, 0.80) | 0.72 (0.69, 0.76) | 0.77 (0.74, 0.80) | **9.6E-05** | 9.3E-01 | **1.4E-04** |
| 0-5 | 10813 (309/10504) | 0.74 (0.71, 0.77) | 0.71 (0.68, 0.74) | 0.75 (0.72, 0.78) | 7.4E-03 | 4.9E-01 | **1.2E-03** |
| **GE Tissue Equalization** | | | | | | | |
| 0-1 | 24525 (183/24342) | 0.81 (0.78, 0.84) | 0.87 (0.85, 0.90) | 0.86 (0.83, 0.89) | **1.1E-05** | **8.3E-05** | 3.9E-01 |
| 0-2 | 23545 (304/23241) | 0.74 (0.71, 0.77) | 0.80 (0.77, 0.82) | 0.79 (0.76, 0.82) | **4.4E-06** | **9.5E-05** | 4.6E-01 |
| 0-3 | 22791 (390/22401) | 0.74 (0.71, 0.76) | 0.78 (0.76, 0.81) | 0.77 (0.75, 0.80) | **9.9E-06** | **5.7E-04** | 3.3E-01 |
| 0-4 | 22129 (492/21637) | 0.71 (0.69, 0.74) | 0.75 (0.72, 0.77) | 0.74 (0.72, 0.76) | **1.2E-03** | 1.7E-02 | 4.2E-01 |
| 0-5 | 21463 (571/20892) | 0.71 (0.69, 0.74) | 0.74 (0.72, 0.76) | 0.73 (0.71, 0.75) | 1.3E-02 | 1.0E-01 | 3.7E-01 |

**Supplementary Table 1.** Pairwise comparison of AUC performance across Native, Finetuned, and Device Invariant models for the cumulative risk score definition. For each year horizon and mammography device, the number of examinations (cases/controls) and AUC (95% CI) are reported for each model. Significant results based on Bonferroni-adjusted threshold are in bold face (DeLong P-Value $< 0.0056$).

| Annual Risk Time Interval | Number of cases/controls | Native Mirai AUC (95% CI) | TE Finetuned AUC (95% CI) | Device Invariant AUC (95% CI) | Finetuned vs. Native (p-value) | Device Invariant vs. Native (p-value) | Device Invariant vs. Finetuned (p-value) |
|---|---|---|---|---|---|---|---|
| **Hologic** | | | | | | | |
| 0-1 | 115774 (1037/114737) | 0.87 (0.85, 0.88) | 0.73 (0.71, 0.74) | 0.87 (0.86, 0.89) | **9.7E-68** | 1.6E-01 | **1.8E-74** |
| 1-2 | 112006 (457/111549) | 0.71 (0.68, 0.73) | 0.60 (0.57, 0.62) | 0.70 (0.68, 0.73) | **4.2E-17** | 4.7E-01 | **2.4E-15** |
| 2-3 | 108629 (494/108135) | 0.69 (0.67, 0.72) | 0.58 (0.55, 0.61) | 0.67 (0.64, 0.69) | **3.7E-18** | **3.9E-03** | **7.9E-12** |
| 3-4 | 104406 (434/103972) | 0.66 (0.64, 0.69) | 0.58 (0.55, 0.60) | 0.66 (0.64, 0.69) | **4.4E-12** | 6.9E-01 | **1.3E-10** |
| 4-5 | 99817 (459/99358) | 0.63 (0.61, 0.66) | 0.55 (0.53, 0.58) | 0.62 (0.59, 0.64) | **7.8E-10** | 9.3E-02 | **5.7E-06** |
| **GE Premium View** | | | | | | | |
| 0-1 | 11866 (103/11763) | 0.89 (0.86, 0.92) | 0.86 (0.82, 0.89) | 0.90 (0.87, 0.94) | 3.8E-02 | 4.3E-01 | 1.6E-02 |
| 1-2 | 11521 (54/11467) | 0.70 (0.62, 0.77) | 0.64 (0.56, 0.71) | 0.74 (0.67, 0.80) | 3.7E-02 | 4.6E-02 | **7.2E-04** |
| 2-3 | 11236 (55/11181) | 0.71 (0.64, 0.79) | 0.61 (0.54, 0.69) | 0.65 (0.58, 0.72) | **3.1E-04** | **4.9E-04** | 2.0E-01 |
| 3-4 | 10893 (45/10848) | 0.68 (0.60, 0.76) | 0.63 (0.54, 0.71) | 0.66 (0.59, 0.74) | 1.6E-01 | 5.8E-01 | 3.0E-01 |
| 4-5 | 10556 (52/10504) | 0.61 (0.54, 0.68) | 0.65 (0.58, 0.73) | 0.64 (0.57, 0.72) | 2.0E-01 | 1.7E-01 | 7.6E-01 |
| **GE Tissue Equalization** | | | | | | | |
| 0-1 | 24525 (183/24342) | 0.81 (0.78, 0.84) | 0.87 (0.85, 0.90) | 0.86 (0.83, 0.89) | **1.1E-05** | **8.3E-05** | 3.9E-01 |
| 1-2 | 23362 (121/23241) | 0.64 (0.59, 0.70) | 0.68 (0.63, 0.73) | 0.68 (0.63, 0.73) | 5.2E-02 | 5.7E-02 | 9.8E-01 |
| 2-3 | 22487 (86/22401) | 0.71 (0.66, 0.76) | 0.73 (0.68, 0.78) | 0.72 (0.67, 0.77) | 3.9E-01 | 8.2E-01 | 5.2E-01 |
| 3-4 | 21739 (102/21637) | 0.63 (0.58, 0.69) | 0.60 (0.55, 0.65) | 0.61 (0.55, 0.66) | 1.2E-01 | 2.5E-01 | 7.7E-01 |
| 4-5 | 20971 (79/20892) | 0.70 (0.64, 0.75) | 0.67 (0.61, 0.73) | 0.66 (0.60, 0.72) | 3.0E-01 | 1.5E-01 | 7.5E-01 |

**Supplementary Table 2.** Pairwise comparison of AUC performance across Native, Finetuned, and Device Invariant models for the annual risk score definition. For each year horizon and mammography device, the number of examinations (cases/controls) and AUC (95% CI) are reported for each model. Significant results based on Bonferroni-adjusted threshold are in bold face (DeLong P-Value < 0.0056).

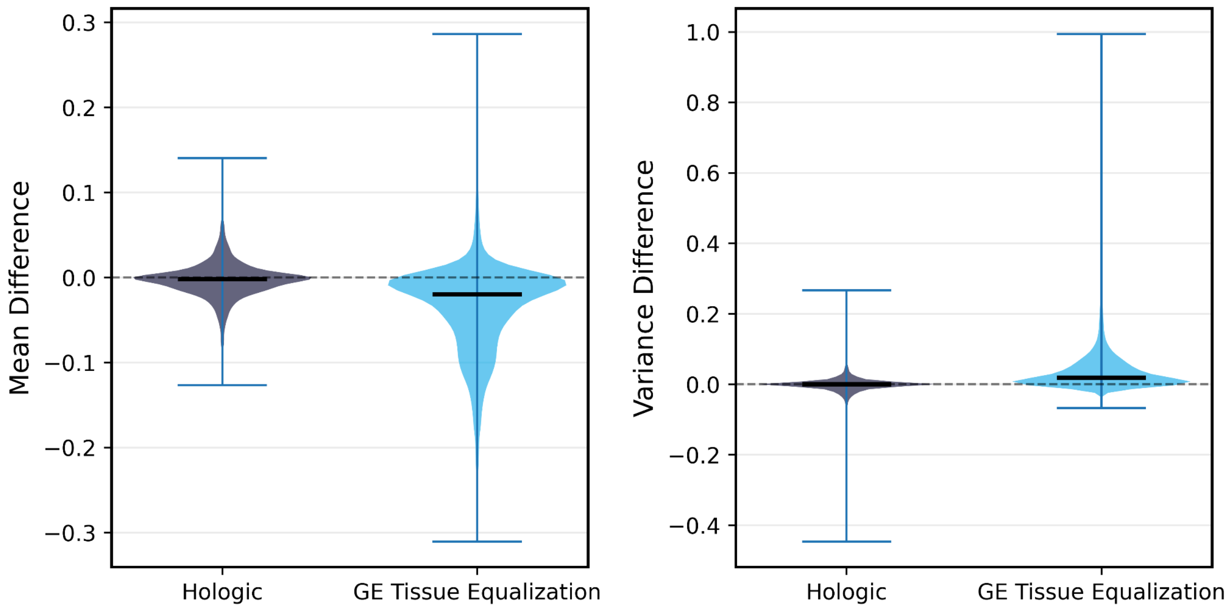


**Supplementary Figure 2.** Violin plot of the distribution of batch normalization activation statistic differences across all Mirai encoder layers and channels, comparing pretrained Mirai running statistics against observed batch statistics from 2,000 randomly sampled Hologic and GE Tissue Equalization mammograms. Each data point represents the difference between the pretrained running statistic and the TE fine-tuning batch statistic for a single filter channel within a single BN layer.
